# An Integrative Network Approach for Longitudinal Stratification in Parkinson’s Disease

**DOI:** 10.1101/2024.01.25.24301595

**Authors:** Barry Ryan, Riccardo E. Marioni, T. Ian Simpson

**Affiliations:** School of Informatics, University of Edinburgh, 10 Crichton Street, EH8 9AB, Edinburgh, UK; Centre for Genomic and Experimental Medicine, Institute of Genetics and Cancer, University of Edinburgh, Crewe Road South, EH4 2XU, Edinburgh, UK

**Keywords:** parkinson’s disease, multi-omics integration, graph neural network, longitudinal

## Abstract

Parkinson’s Disease (PD) is a neurodegenerative disorder characterized by motor symptoms resulting from the loss of dopamine-producing neurons in the brain. Currently, there is no cure for the disease which is in part due to the heterogeneity in patient symptoms, trajectories and manifestations. There is a known genetic component of PD and genomic datasets have helped to uncover some aspects of the disease. Understanding the longitudinal variability of PD is essential as it has been theorised that there are different triggers and underlying disease mechanisms at different points during disease progression. In this paper, we perform longitudinal and cross-sectional experiments to identify which data modalities or combinations of modalities are informative at different time points. We use clinical, genomic, and proteomic data from the Parkinson’s Progression Markers Initiative. We validate the importance of flexible data integration by highlighting the varying combinations of data modalities for optimal stratification at different disease stages in idiopathic PD. We show there is a shared signal in the DNAm signatures of participants with a mutation in a causal gene of PD and participants with idiopathic PD. We also show that integration of SNPs and DNAm data modalities has potential for use as an early diagnostic tool for individuals with a genetic cause of PD.

## I. Introduction

Parkinson’s Disease (PD) is a heterogenous, progressive, multisystem neurological disorder that affects the nervous system. It is most commonly characterised by a range of motor symptoms, primarily involving difficulties with movement, however a wide variety of non-motor symptoms also exist. PD has a complex pathophysiology, but these disease pathways culminate in the gradual death of neuronal cells, causing a deficit in dopamine [27].

One notable aspect of PD is the variability between individuals with the disease. PD is characterised by core motor syndromes of tremor, rigidity, bradykinesia and postural instability. The onset, trajectory and experience of these symptoms among people varies significantly. Genetic mutations in individuals account for approximately 30% of cases, however not everyone with a mutation will develop the disease [10]. The trajectory of the disease among patients is highly variable, with some experiencing a rapid progression to disability and others following a relatively benign course [23]. Whether an individual develops all motor and any non-motor symptoms can vary too. While PD medications do not cure the disease, they do help with some of the day-to-day motor symptoms, however the time period for which they are effective varies between patients also [3].

Identification of mutations in single genes have aided the understanding of PD. For example, specific variants in the *LRRK2, GBA*, and *PINK1* genes are associated with PD [3]. This motivates the use of omic measures for uncovering novel insights into the pathology of PD. Omic data modalities capture genetic and/or biomolecular profiles; analyses of these data has resulted in many novel findings in PD. Craig et al. (2021) found early alterations between the gene expression of PD patients and healthy individuals [2]. Similarly, Kern et al. (2021) found that non coding RNA’s can have diagnostic and prognostic power in PD individuals [8]. Recent Genome Wide Association Studies of PD have had conflicting results. Walters et al. (2023) found no genome wide significant loci for PD in the China Kandoori Biobank with a population of 105,408 Chinese individuals [26]. Conversely, in a population of 2478 Chinese individuals, Pan et al. (2023) found 19 associations with PD including genome wide significant loci in *LRRK2, SNCA*, and *GBA* [18]. Currently, there is no known exogenous or genetic trigger for PD that causally results in the loss of dopaminergic cells.

It has been hypothesised that the disease mechanisms of PD change over time and that treatment needs to account for disease stage as well as individual molecular and disease phenotypes [27]. Longitudinal variability poses significant challenges in both the biological understanding and treatment of PD. This heterogeneity necessitates a flexible approach that can incorporate multiple sources of information at a given stage of PD. The Parkinson’s Progression Markers Initiative (PPMI) was created for this reason. It consists of longitudinal clinical, genomic, and imaging data from over 900 PD cases, 800 Prodromal (cases without a clinical diagnosis for PD, but early indicators that they will go on to develop it) and 230 Healthy Controls.

We propose a flexible integrative approach using a network taxonomy that can incorporate many aspects of the PPMI dataset, most notably the longitudinal component. Variability in disease motivates an individualised approach to disease management. We utilise a patient similarity measure to identify patients who have similar molecular, epigenetic, and demographic disease characteristics. We hypothesise that integrating many sources of complementary information can unravel many of the unknown aspects of PD. By combining a patient-focused approach with multiple sources of information, we hope to learn what differentiates PD patients from those who have early signatures of the disease and healthy controls.

Multi-modal approaches have achieved good prediction accuracy using the PPMI dataset. Chan et al. (2022) achieve perfect disease stratification using a model which incorporated multiple omic and image datasets [1]. A review by Gerraty et al. (2023), on multi-modal integration approaches in the PPMI dataset, found that clinical and neuroimaging datasets were the most commonly used modalities [5]. They further identified that few machine learning focused papers use the longitudinal structure of the PPMI study. A possible reason for this is due to restricted patient coverage when incorporating image data. Chan et al. identify that the dataset they utilised is small and heavily skewed to PD patients [1]. Given the time-consuming nature and expense of collecting image data, this is not surprising.

In this analysis, we integrate omic datasets such as Messenger RNA expression (mRNA) and Single Nucleotide Polymorphisms (SNPs) with clinical and proteomic information using a flexible network taxonomy that allows retention of the maximum number of patients in the analysis. We represent the integrated modalities as a Patient Similarity Network (PSN) and use an Graph Neural Network (GNN) architecture for disease stratification. We group patients into those with a mutation in a known causal gene for PD, those who have a sporadic onset of the disease, and finally a combination of both. In each case we attempt to classify individuals as either having PD, being prodromal, or a healthy control. We perform experiments cross-sectionally across 4 time points over the course of the first three years of a patient’s disease post diagnosis. We assess the best combination of modalities at each time point and contrast the findings between the three groups. Finally, we re-run the analysis on a subset of genetic PD patients who have data across all time points, with a model trained at each time point. The goal of this experiment is to identify whether the disease signatures we identify change over the first three years of the disease by assessing if the learnt biological signals remain consistent across the 4 time points.

## II. Methods

### Multi-Omic Graph Diagnosis (MOGDx)

MOGDx is a flexible tool to integrate multiple omic measures and perform classification tasks. This approach uses a network taxonomy to combine patient similarity matrices into a single network and perform node classification using a Graph Convolutional Network (GCN). The performance of MOGDx was benchmarked on cancer data and achieved state-of-the-art performance compared to similar research [22].

MOGDx can integrate any number of modalities. This includes omic measures as well as any other modalities of interest, such as clinical descriptors. A single Patient Similarity Network (PSN) is built per modality. The most informative features of each modality are used to inform the similarity metric with similarity between patients calculated using Pearson correlation, where suitable, otherwise Euclidean distance. As is common practice, all patient information is used to construct the network, with train, validation and test labels created during the training phase of the GCN-MME [11], [30]. In this approach, non-informative features are discarded to reduce the complexity of the similarity calculation and to discourage uniform similarity scores for modalities such as DNAm which will have large number of redundant or similar features. Each PSN is constructed using the k nearest neighbours algorithm, and the Similarity Network Fusion (SNF) algorithm is used to combine individual PSN’s into a single network.

The fused PSN and the modalities are input into the Graph Convolutional Network with Mulit-Modal Encoder, shown in Figure S1. Each modality is compressed using a two layer encoder. The first layer of the encoder is of fixed length, with the second layer being tuned to each modality by performing a hyperparameter search. Median imputation is performed on the second layer of each encoder to retain patients if they are missing from that modality. The compressed encoded layer of each modality is then decompressed to a shared latent space using mean pooling. Similar encoder architectures have been established in other works [28], [29]. This shared latent space corresponds to the patient node features, which are combined with the PSN and input into the GCN for classification. The GCN-MME is trained under the semi-supervised setting for GNN outlined by Hamilton (2020) [7]. For a detailed description of the MOGDx architecture, please refer to Ryan et al. 2023 [22].

MOGDx is a suitable tool to perform analysis on the PPMI dataset due to its flexibility. It can integrate any number of modalities, whilst simultaneously retaining the maximum number of patients possible, in contrast to other existing methodologies. As discussed by Chan et al. (2022) and as per Figure 1, there are relatively few healthy control participants [1]. Not every participant will be present in each modality at each time point. In order to avail of the full PPMI dataset, a method which can incorporate the maximum number of participants is required. MOGDx achieves this by utilising SNF and imputation to retain patient nodes. Moreover, including patients missing in one or more modalities does not result in a large degradation in performance [22]. MOGDx provides a high level of interpretability. Due to the flexibility of integration, ablation experiments can be performed to identify the most predictive modalities. As the most informative features are extracted in the MOGDx pipeline, these features can be further analysed to identify important pathways, traits or interactions of the target application.

**Fig. 1.**
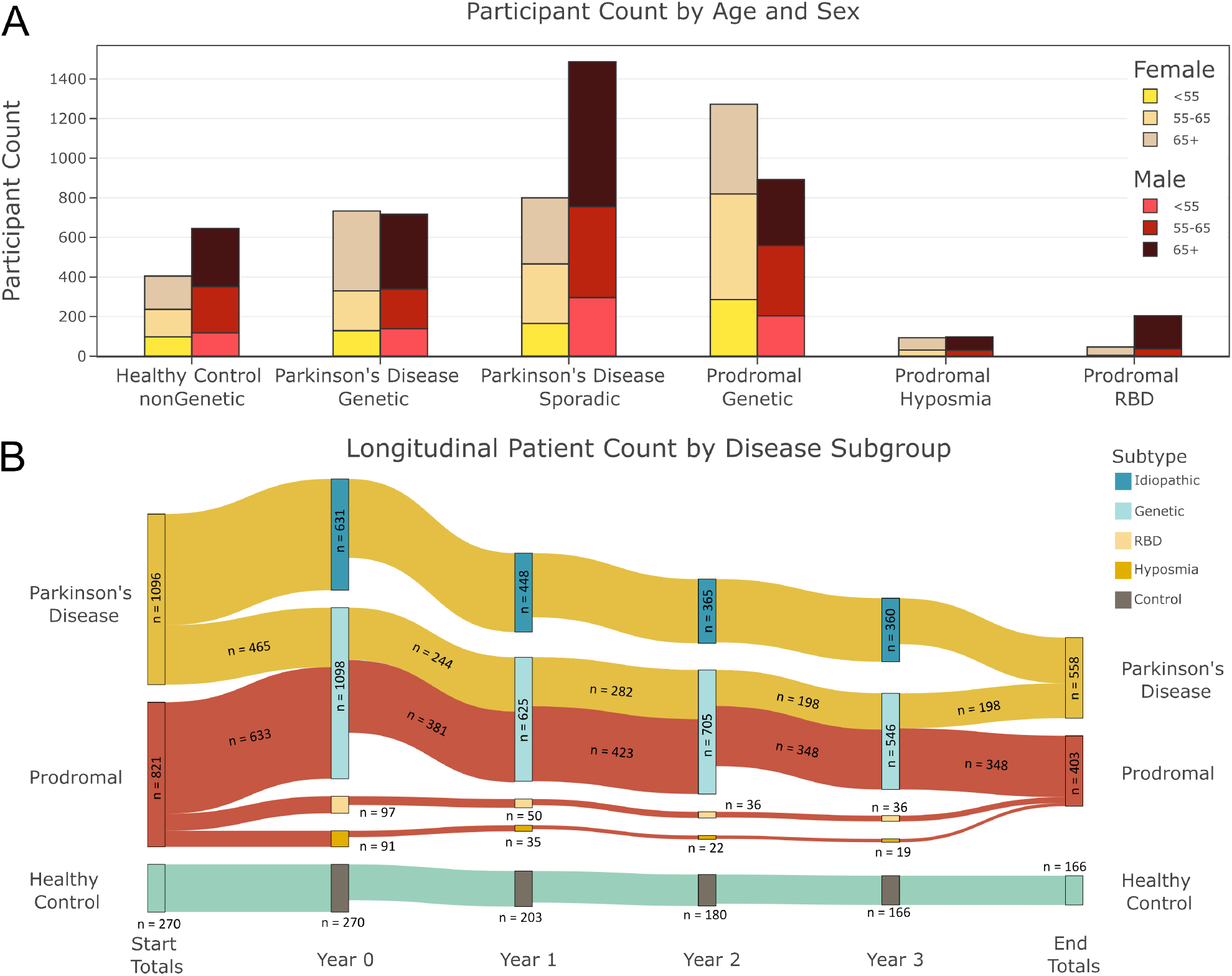
**A Participant Count by Age and Sex —** The number of participants in each disease subgroup broken down by age and sex are shown. PD idiopathic and Genetic Prodromal are the two largest cohort subgroups. The majority of participants are older than 55 years, with a slightly larger male majority. Relatively, there are very few PL participants who do not have genetic predisposition. **B Longitudinal Participant Count by Disease Subroup —** The flow of participant availability over the first four years in PD, PL and HC participants in the PPMI study. The number of participants available decays in all subgroups over time.

### PPMI Dataset

Data was obtained from the PPMI [14]. The modalities analysed and number of features per modality at year 0 are summarised in Table I. All other time points are included in Tables S1-S3 in the supplementary. In total, 5988 samples from 2188 participants were included in the analysis, as per Table S4. Patient characteristics and the participant sample availability over time are shown in Figure 1. Participants in the analysis were identified as Parkinson’s Disease (PD), Prodromal (PL) or Healthy Control (HC) participant.

**TABLE I.**
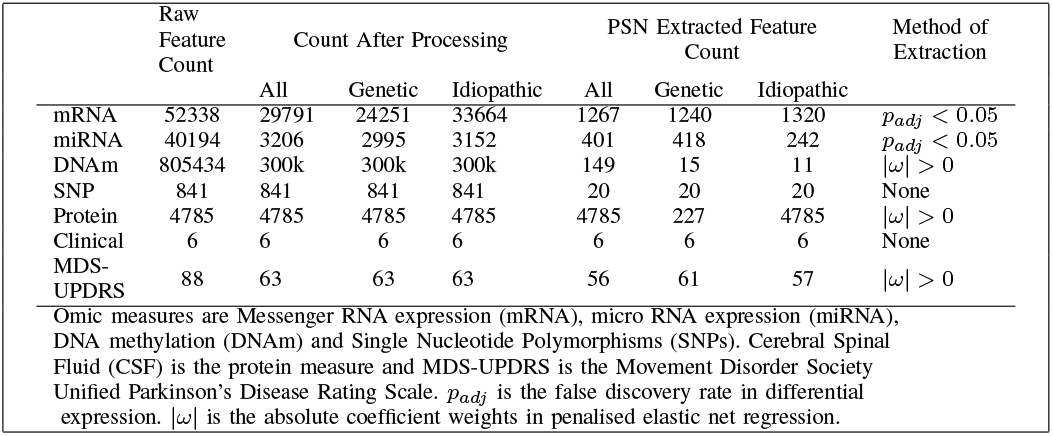
Breakdown of Modality Features in PPMI Dataset at Year 0.

Four genomic measures, shown in Table I, were generated from whole-blood samples and analysed. Each modality was processed in a complimentary bioinformatics pipeline if available. For example, DNAm was normalised using the wateRmelon package in R [20]. See Ryan et al. (2023) for more detail on how specific genomic modalities were handled [22]. In general, processing included the removal of uninformative or missing features, normalisation, imputation of missing values and conversion from categorical features to numerical features. The top 300k most variable CpG sites were retained to allow computation on this dataset to fit into memory. For the same reason, a Principal Component Analysis (PCA) was performed on the SNPs dataset to reduce the dimensionality of the dataset, and the first 20 PC’s were retained.

Genomic datasets were supplemented with additional measures of 1472 CSF markers extracted from participants and clinical descriptors. Clinical descriptors included individual phenotypes of age, sex and years of education; These were supplemented with measures for smoking, alcohol and BMI generated from DNAm profiles [15]. These DNAm profiles were derived from models trained on up to 5087 individuals in a national study in Scotland and tested on two separate cohorts also based in Scotland [15]. The MDS-UPDRS by Goetz et al. (2008) is a measure of disease severity in those with PD and PL [6]. This scale combines measures relating to both motor and non-motor symptoms of PD. It consists of both self-assessment and clinical assessments and is a proxy of disease stratification [6]. It was used as a baseline comparative model to identify if the biological signal for PD found in the blood is stronger than clinical assessment using MOGDx. These modalities were similarly processed for feature removal, conversion and normalisation.

Pairwise linear regression between the three classes was performed using the DESeq2 package in R to obtain differential gene expression transcripts [12]. For non gene expression modalities, penalised elastic net regression was performed using the glmnet package in R [25]. Differentially expressed genes with a statistically significant FDR (*p*_*adj*_ < 0.05) and logistic regression coefficients with an absolute weight greater than zero were used for feature selection prior to the similarity calculation. This discourages uniform similarity scores for modalities such as DNAm which will have large number of redundant or similar features. The number of informative features is dependent on the subgroup being analysed. If no informative features were found, all features were retained. Further information on the experiments is included below, with the feature counts summarised in Table I.

Participant samples have been broken down by sex, age, subgroup and time point in Figure 1. The time points cover the first three years of the disease in the PD cohort. The first time point (labelled year 0) corresponds to participants with PD who have had a diagnosis for less than 2 years, have not begun taking any PD medication and are not expected to require PD medication for at least 6 months [14]. Those in the genetic subgroup of PD have a mutation in one of three genes: *LRRK2, SNCA* or *GBA*. Idiopathic individuals do not have mutations in any of these three genes. PL participants have been identified as being of high risk for the disease, but have not yet met a clinical threshold for diagnosis. The first time point, year 0, in this cohort corresponds to their enrolment in the study. The genetic subset of this group also have a mutation in one of the three aforementioned genes as aligned with the genetic PD subgroup. As per Figure 1 A, the PL participants in the genetic subgroup far outnumber the participants in the Rapid eye movement Behaviour Disorder (RBD) and hyposmia subgroups. Participants in these groups have one of two non-motor symptoms associated with PD. RBD is a sleep disorder which has been identified as an early indicator for the disease, and hyposmia is a smell disorder which is an early indicator of PD [13], [21]. The HC arm of this analysis have been screened to ensure they did not meet the criteria for either PD or PL. As with PL, their first time point, year 0, aligns with their enrolment in the PPMI study. PD idiopathic and PL genetic are the two most prevalent subgroups in the dataset. The vast majority of participants are aged 55 years or older, and the mean age of all participants is 63 years. As identified in Chan et al. (2022) there are fewer HCs compared to PD and PL however the numbers presented in both Figure 1 A and Table S4 show higher counts compared to their analysis which was subset to participants who had image data available [1]. A distinguishing factor of this analysis is the utilisation of the longitudinal data in the PPMI dataset. Figure 1 B shows the flow of data availability over time. It is split by clinical diagnosis of PD, PL or HC and is further divided at each time point by disease subgroup. It shows that, over time, the number of participants decreases across all diagnoses and subgroups. This is due to participant dropout (n = 401), missing samples for a participant at a time point (see Table S5) or the transition of a PL patient to a clinical diagnosis for PD (n = 33). A summary of the criteria for participant stratification and disease subgroups are summarised in Figure S2.

#### A. Design of Data Analysis

In this analysis, we perform cross-sectional experiments at 4 time points over three years, as well as longitudinal experiments on participants who have a mutation in one of the three causal genes of PD. In all cross-sectional experiments we classify whether participants have PD, are PL or are a HC. We perform these cross-sectional experiments on all participants, regardless of their subgroup. Similarly, we perform the experiments on two subsets based on participants’ subgroup. The first subset, referred to as genetic, includes PD and PL participants in the genetic subgroup. The other subset, referred to as idiopathic, includes all participants in the idiopathic PD, RBD and hyposmia subgroups. HC participants are included in both subsets as a control. We use a brute-force approach, testing all combinations of modalities in each experiment to identify the modalities at each time point with the highest accuracies and F1 scores.

In the longitudinal analysis, we re-perform the best performing cross-sectional experiment on the genetic PD and PL subgroups, with exact numbers shown in Table S6. Once again, HC are included as a control. This analysis includes participants who are present at each time point in at least one of the included modalities. The best performing cross-sectional experiment was determined by averaging the F1 scores of each model across all time points. For this analysis, only the optimal combination of modalities which maximised both accuracy and patient retention was analysed. The longitudinal experiments comprise 4 cross-sectional experiments where MOGDx is trained and tested at each time point and 12 longitudinal experiments where each model is tested on the unseen networks and omics from the alternative time points. Networks are constructed using only the features identified at that model’s time point. For example, when testing the year 3 network at year 1, the network being tested is reconstructed only using the features identified at year 1. In this manner, each of the 4 models are tested on a completely unseen test set of the same patients but from other time-points. This is undertaken to assess if the biological signal learnt at one time point is present at other time points.

## III. Results

### Performance & Evaluation

The performance metrics used to compare the classification performance of MOGDx were accuracy, F1 score and improvement in accuracy. The F1 score was calculated by the mean F1 score of each class, weighted by the size of that class. Improvement in accuracy is a metric used to compare how much the accuracy improved compared to a baseline model. In this case, the baseline model is a simple model which predicts the most common class. Stratified k-fold cross validation was performed with 5 randomly generated splits to obtain the mean and standard deviation metrics reported. Within each split, the set was further randomly split into training and validation sets to produce an overall train/validation/test split of 68%/12%/20% respectively.

#### A. An integrative approach is optimal when classifying individuals with PD over time

The results from the cross-sectional experiments, shown in Table II and Figure 2, highlight the power of a flexible integrative approach when classifying participants in the PPMI dataset with PD. The flexibility of the approach allows us to test all modalities individually, as well as all combinations of integrated modalities at each time point. As a result, all 6 modalities are included in at least one experiment. This is further evident in Figure S3, which shows that an integrated approach is preferred in 13 of the top 15 best performing models averaged across all time points of the three groups. In Table II there are only two experiments, years 1 and 3 with all participants (genetic + idiopathic), which do not integrate modalities for optimal performance. In Figure 2, the three worst performing models are all individual modalities, whereas the best model in the genetic and idiopathic subgroups integrate two modalities. DNAm performs best individually when predicting all participants. The improvement in accuracy of these DNAm models, at most time points in Figure 2, is lower than the combined modalities in the other subgroups. There is an increase in accuracy compared to the worst performing modality, miRNA, but it does not match or improve on the baseline MDS-UPDRS assessment. Only the genetic subgroup achieves an improvement in accuracy greater than the MDS-UPDRS assessment. This could motivate the use of these modalities for early disease diagnosis, as motivated below. The combination of CSF and DNAm in the idiopathic subgroup shows promising performance, particularly at year 2. The MDS-UPDRS is an accurate baseline to compare to, given it consists of clinical assessment scores of both motor and non-motor symptoms [6]. Thus, the results show encouraging performance when integrating combinations of modalities in subgroups of PD.

**TABLE II.** Cross-Sectional performance of MOGDx in different subgroup experiments.

|  |  | Modalities | Number of Participants | Accuracy | F1 score | Improvement in Accuracy |
| --- | --- | --- | --- | --- | --- | --- |
| Genetic + Idiopathic (All) | Year 0 | DNAm + SNP + mRNA + miRNA | 1515 | $0.630 \pm 0.019$ | $0.665 \pm 0.017$ | $0.110 \pm 0.018$ |
| | Year 1 | DNAm | 548 | $0.624 \pm 0.020$ | $0.667 \pm 0.032$ | $0.111 \pm 0.02$ |
| | Year 2 | Clinical + DNAm | 542 | $0.694 \pm 0.037$ | $0.717 \pm 0.034$ | $0.166 \pm 0.037$ |
| | Year 3 | DNAm | 493 | $0.712 \pm 0.018$ | $0.699 \pm 0.048$ | $0.146 \pm 0.018$ |
| Genetic | Year 0 | DNAm + SNP | 489 | $0.789 \pm 0.036$ | $0.753 \pm 0.04$ | $0.419 \pm 0.036$ |
| | Year 1 | DNAm + SNP | 443 | $0.867 \pm 0.018$ | $0.835 \pm 0.02$ | $0.472 \pm 0.018$ |
| | Year 2 | DNAm + SNP | 432 | $0.866 \pm 0.031$ | $0.837 \pm 0.032$ | $0.477 \pm 0.031$ |
| | Year 3 | DNAm + SNP | 365 | $0.841 \pm 0.034$ | $0.811 \pm 0.038$ | $0.403 \pm 0.034$ |
| Idiopathic | Year 0 | SNP + miRNA | 667 | $0.681 \pm 0.031$ | $0.752 \pm 0.008$ | $0.069 \pm 0.031$ |
| | Year 1 | CSF + DNAm + SNP | 582 | $0.720 \pm 0.039$ | $0.776 \pm 0.035$ | $0.122 \pm 0.039$ |
| | Year 2 | CSF + Clinical + DNAm | 399 | $0.805 \pm 0.022$ | $0.770 \pm 0.022$ | $0.246 \pm 0.022$ |
| | Year 3 | CSF + DNAm | 360 | $0.764 \pm 0.022$ | $0.721 \pm 0.021$ | $0.183 \pm 0.022$ |

**Fig. 2.**
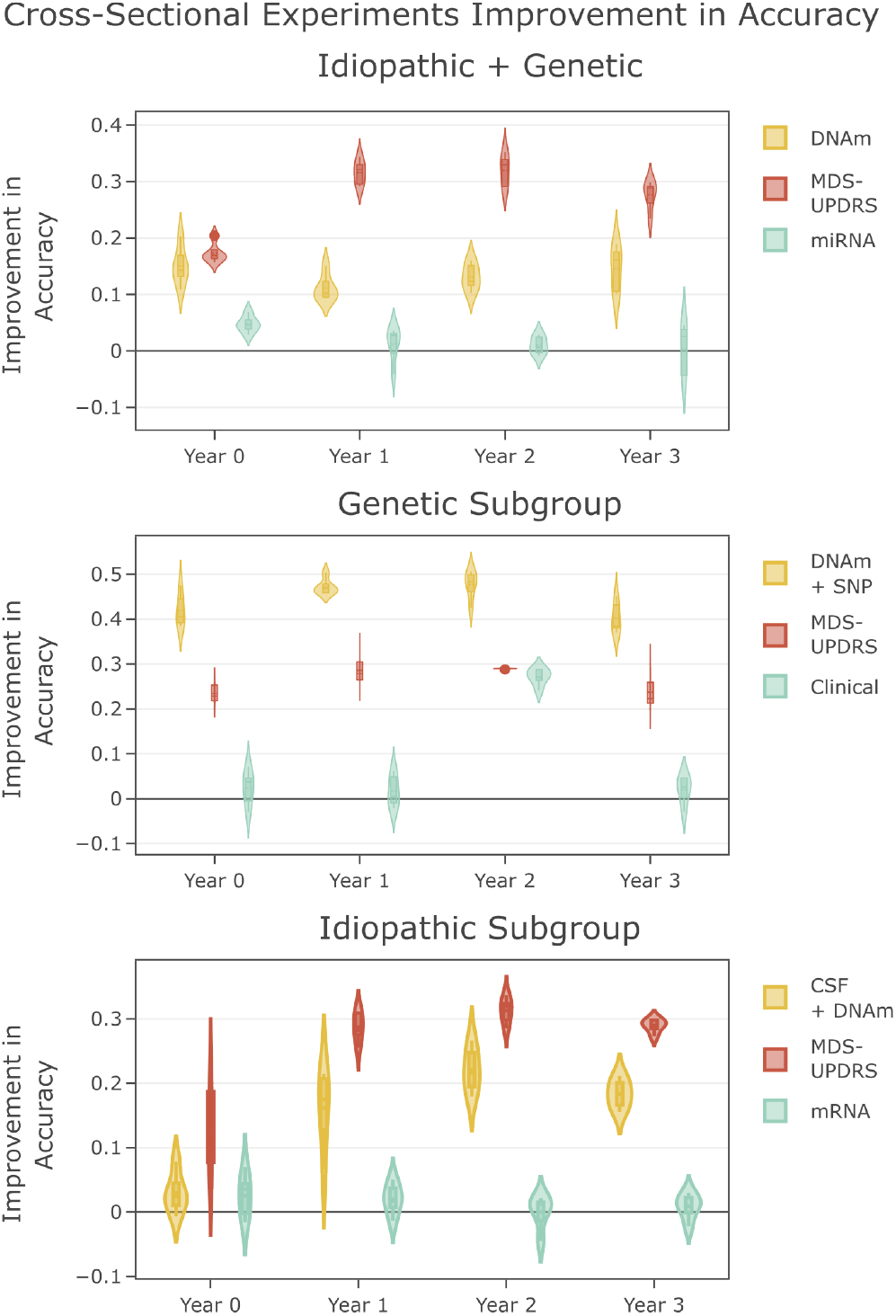
Modality Integration Performance of Best, Worst and Baseline (MDS-UPDRS) Models. **A — Idiopathic + Genetic** Optimal performance, using DNAm, does not improve on baseline MDS-UPDRS assessment but is better than worst performing modality miRNA **B — Genetic** Integrating DNAm and SNPs for participants with a genetic predisposition for PD performs better than the MDS-UPDRS assessment and worst performing modality **C Idiopathic —** Integrating CSF with DNAm improves on the worst performing modality, mRNA, but has worse performance compared to the MDS-UPDRS assessment.

#### B. Flexibility in integration of modalities facilitates a biological signal for PD to be learnt from whole-blood samples and protein markers in PPMI study participants

Table II highlights the importance of flexibility when integrating different modalities. There is an improvement in accuracy compared to a model which predicts the most common class in all experiments. This improvement increases with time, indicating an increased biological signal for PD as the disease progresses. In both genetic and idiopathic subgroups, years 1 and 2 are the most predictive time points. This could indicate that these time points are capturing both early and late signatures of PD. This is particularly evident in the idiopathic subgroup, where there is a change in predictive modalities over time, with common modalities early and late in the disease. The genetic SNPs modality is predictive early, whereas protein CSF markers along with DNAm are more prominent in later stages. This supports the work of Wüllner et al. (2023) who found that there may be different disease mechanisms at different stages of PD [27]. The caveat is that the prediction accuracy at year 0 in this subgroup is low.

Conversely, in the genetic subgroup, the modalities which are most predictive do not change with time. Unsurprisingly, the SNPs modality is included across all time points in the genetic subgroup experiments. SNPs are a fixed description of participant genetic information [4]. Thus, this dataset can clearly distinguish HCs from PD and PL individuals who have a mutation in a known causal gene for PD. It does not differentiate between PD and PL participants, thus indicating that this differentiation is learnt from another modality, in this case DNAm. Whether the biological signal learnt changes over time requires further work to understand the drivers of variability in DNAm at each time point. The integration of these two modalities outperforms the MDS-UPDRS baseline, highlighting the predictive power of using whole-blood samples to extract omic information relating to a neurological disease. In summary, we found a strong disease signal both early, but particularly late, in the blood of individuals with a genetic predisposition for PD, despite it being a neurological disorder.

#### C. There are possible similarities in the DNAm signatures of idiopathic participants and participants who have a genetic predisposition for PD

There is a clear genetic driver in participants who have a mutation in a causal gene of PD. As per Table 2, the genetic subgroup achieves the highest accuracies, F1 scores and improvements in accuracy at all time points. As included participants have a mutation in one of the *LRRK2, GBA* or *SNCA* genes, the genetic influence on their disease is far more prominent and can be distinguished with high classification accuracy using genomic data. This highlights the homogeneity between participants in this subgroup and the power of using a patient similarity approach for tasks of this nature. The idiopathic subgroup contains participants with unknown causes of PD or, in the case of participants labelled PL, have an early indication of developing the disease. This group has no known genetic association with PD, therefore it is unsurprising that the accuracies achieved when integrating genomic data is lower. It is still possible that there is a genetic cause of PD in this cohort, however there are likely numerous signatures which are too diverse for a signal to be found.

This makes the idiopathic cohort very heterogenous. Despite these significant differences between participant subgroups, most experiments include DNAm as a predictive modality, with the idiopathic subgroup at year 0 being the only experiment where it is not included. Given this prominence, it indicates the presence of epigenetic modifications between PD, PL and HC participants. When considering all participants (genetic + idiopathic), there is a mix of a homogeneous and a heterogeneous group, which makes learning very difficult. Despite this, there is a robust improvement in accuracy between 10% and 20% across all time points, as per Figure 2. This suggests that there may be a shared signal between the two subgroups in the DNAm modality. A possible explanation for the decreased performance compared to the two subgroups is that the additional information added by other integrated modalities is not shared between the two subgroups. In order to confirm if the signal being learnt is similar, more research needs to be conducted to identify the common discriminating DNAm features, however our results suggests common signatures in the DNAm of genetic and idiopathic PD participants.

#### D. An integrative model trained at a late disease stage could form a viable early diagnostic tool for predicting individuals with PD who have a genetic predisposition for the disease

In the cross-sectional experiments, we show the metrics for classifying participants with a mutation in a causal gene of PD to be very promising. The combination of DNAm and SNPs achieves a consistently high accuracy, F1 score and improvement in accuracy. The improvement in accuracy is consistent across all time points, as per Figure 2, highlighting the robustness of the signal learnt. At year 0, participants with PD are in the early stages of their disease. They have had a clinical diagnosis for two years or less, have not begun taking medication and are not expected to be required to take medication for at least 6 months. Despite this, the models are able to discriminate between the three stratification targets. Further research should be conducted to identify if this signal can be learnt prior to diagnosis and motivates the integration of DNAm with SNPs for early PD detection. Longitudinal experiments were performed on a subset of participants from the genetic group who are present in either DNAm or SNPs at each time point. These experiments were designed to identify the optimal time point to train such a diagnostic tool and if the disease signal learnt early in the disease is present later and vice versa.

Table III shows the results of the longitudinal experiments and clearly highlights that an early PD detection model should be trained later in the disease course. Both the accuracy and F1 scores increase with models which are trained later in the disease course. Optimal performance was observed by the model trained at year 3. Poorest performance was observed by the model trained at year 0, with the performance of models trained at years 1 and 2 being comparable. For simplification of comparison, the metrics reported in Table III, report the accuracy and F1 score achieved when the model classifies all participants included in the experiment. Therefore, only for the models trained and tested at the same time point, 68% of the participants will have been seen by the model in the training set. This accounts for the apparent increase in accuracy relative to the cross-sectional metrics reported in Table II. Despite this, the model trained at year 3 achieves a higher accuracy when tested at year 0 and year 2 compared to the models trained at these time points. While the model trained at year 3 doesn’t improve on the accuracy of the model trained and tested at year 1, it does outperform all models at all other time points, as per Figure S4.

**TABLE III.** Longitudinal Experiments Performance Metrics.

| Accuracy / F1 |  | Time Point Model Tested |  |  |  |
| --- | --- | --- | --- | --- | --- |
|  |  | Year 0 | Year 1 | Year 2 | Year 3 |
| Time | Year 0 | 0.832 / 0.798 | 0.806 / 0.768 | 0.813 / 0.773 | 0.816 / 0.777 |
| Point | Year 1 | 0.839 / 0.815 | 0.911 / 0.886 | 0.845 / 0.811 | 0.845 / 0.814 |
| Model | Year 2 | 0.849 / 0.814 | 0.849 / 0.814 | 0.872 / 0.840 | 0.836 / 0.799 |
| Trained | Year 3 | 0.895 / 0.874 | 0.888 / 0.870 | 0.908 / 0.885 | 0.947 / 0.932 |

Figure 3 shows the accuracy broken down by class for the four models trained at each time point. All models predict the HC class with high accuracy. As mentioned, both PD and PL participants have a genetic risk variant for the disease, thus, the SNPs modality can easily discriminate between them and the HC participants. The main differentiation between the models is their ability to distinguish PD from PL participants. In general, it can be observed from Figure 3 that the accuracy in predicting PD participants decays the further away in time you test the model from when it was trained. This can be observed in Figure 3, both by the sharp gradients of the PD participants when assessing the number of consecutive correct predictions of a model and the decrease in flow accuracy. Conversely, the PL class have much more stable and consistent predictions across all time points. This is evident in Figure 3 with the number of PL participants correctly classified in the flow diagram being less variable over time and the flatter gradients in the consistency of predictions.

**Fig. 3.**
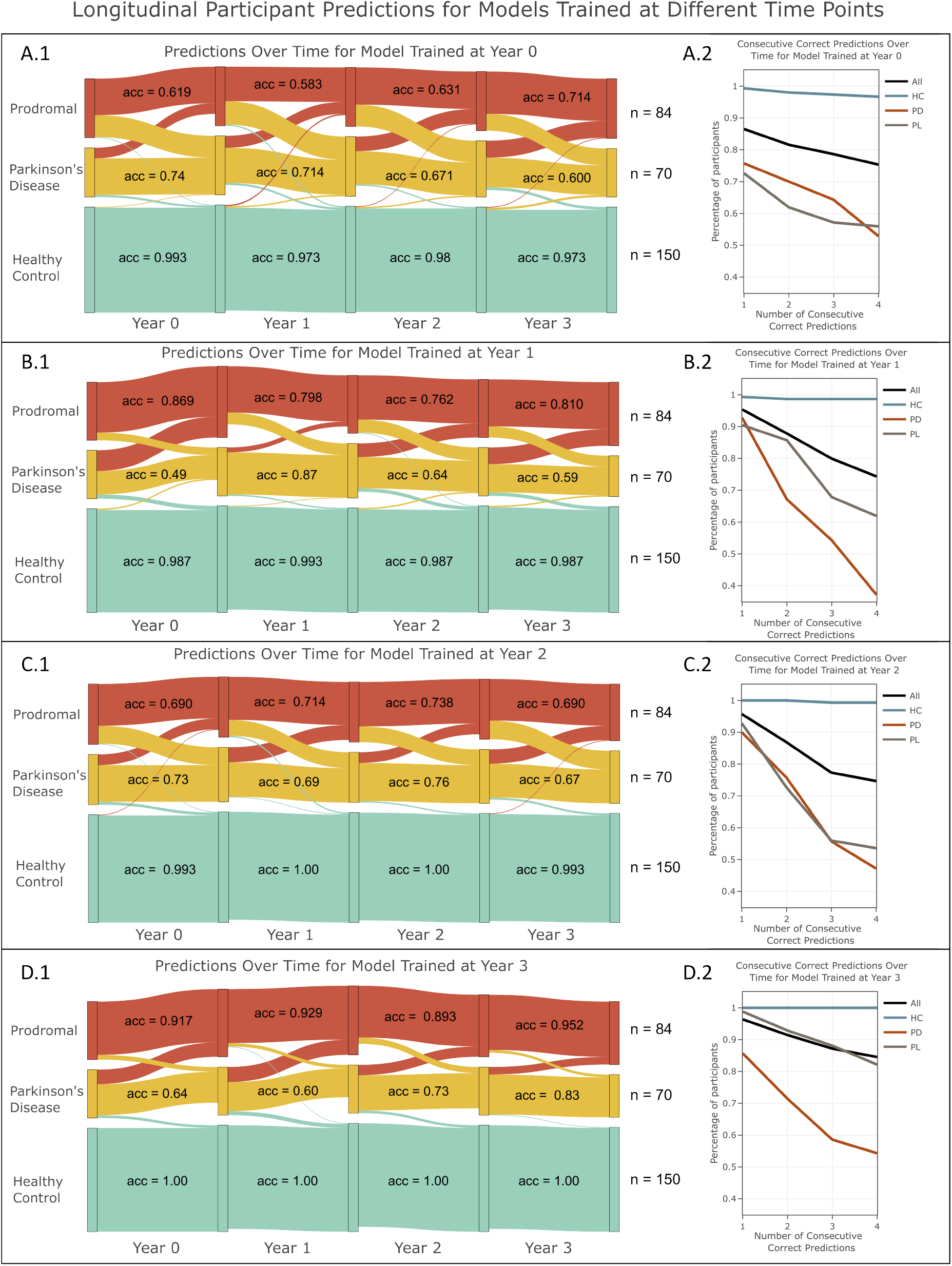
Longitudinal Experiments Participant Stratification. **A — Year 0** This model consistently predicts the same participants as PD, PL and HC which can be observed by the consistent flow of predictions in **A.1** and relatively flat gradients in **A.2. B — Year 1** This model is very accurate when trained and tested at the same time point, but performs poorly when predicting PD participants at other time points. This leads to significant changes in the flow of predictions in **B.1** and sharp gradients in **B.2. C — Year 2** This model achieves good trade off in predicting between PD and PL participants as can be seen by symmetry in **C.1**, but the predictions are not consistent as per the sharp gradients in **C.2. D — Year 3** This is the best performing model. There is good symmetry in predictions in **D.1** and the lines in **D.2** are relatively flat. It does have more difficulty predicting PD participants earlier in the disease course, thus the sharper decline in **D.2**.

In Table 3, we show there is a much stronger signal discriminating PD from PL participants later in the disease course. This finding is expected as the PD participants, on average, will have a more severe disease at year 3 than they will at year 0. What these results therefore show is that by year 3 we have found a very accurate threshold for differentiating PD participants from PL. When we then back-propagate this threshold by testing the model over time, we find that the PL participants maintain a high predictive accuracy, but some PD participants cross this threshold and are misclassified as PL. As stated, differences between these groups can be largely explained by differences in their DNAm. Thus, we can attribute these findings to epigenetic modifications occurring in participants with PD as their disease progresses.

## IV. Discussion

In this paper, we applied an integrative network framework and artificial intelligence to the PPMI dataset. The PPMI dataset is an observational, international study, consisting of multiple data modalities, with the goal of identifying markers of PD to accelerate disease modifying clinical trials [14]. We used clinical, genomic, and proteomic data to include a significant number of patient samples and conducted cross-sectional and longitudinal stratification of participants who have PD, have an early indication of developing PD (Prodromal), or were a Healthy Control.

We found that a flexible integrative approach is optimal when performing disease stratifications for PD. Our models show a strong preference for including multiple modalities. It is clear that there is not sufficient information in any one single modality to accurately capture significant variability in PD at all time points. This highlights the importance of integrating multiple sources of information to capture different components of the heterogeneity in PD. Flexibility is also a key characteristic of this framework. Our approach allows us to test all modalities individually and all combinations of modalities at each time point. This allows us to perform ablation experiments to identify the most informative modalities at each time point. The idiopathic subgroup contains individuals with no known cause of PD. This makes them a very heterogeneous group as there could be a vast number of different disease mechanisms at play, which may not be captured by the clinical, genomic or proteomic data. Our results show improvement in accuracy over a baseline predictive model. The availability of CSF is very informative in this subgroup. Unlike whole blood samples, that can only contain biomolecules that pass through the blood brain barrier, data derived from CSF likely contains a richer biomolecular complement that more closely mimics signatures in the brains of idiopathic PD participants. CSF was included in three experiments in the idiopathic subgroup, despite it only being available in a relatively low number of participants. Unfortunately, only one participant in the genetic subgroup had a CSF sample available. Thus, it is unknown if CSF is informative in the genetic subgroup of the PPMI dataset. As a result, it was not included in any genetic subgroup analyses and its effect was likely obscured in the joint genetic and idiopathic analysis. It is known that CSF is a good marker for PD as multiple CSF measures, in particular CSF *α*-synuclein, are known to be good prognostic measures of PD [19]. The build up of *α*-synuclein is well established in the pathology of PD, particularly later in the disease course, which mirrors our findings of CSF being more predictive later in the analysis [3]. We found that different modalities are informative at different stages of PD in the idiopathic subgroup. This supports the theory, by Wüllner et al. (2023), that the pathology or mechanisms of PD may change over time in this group and highlights the importance of flexibility when integrating different modalities [27].

The strongest metrics were observed when stratifying the genetic subgroup. This group consists of participants who have a mutations in one of three genes, *LRRK2, GBA* or *SNCA*, which are known to be associative with PD [3], [24].

In comparison to the idiopathic group, this genetic group can be considered homogenous as there is a clear genetic driver to their disease. Unsurprisingly, this is reflected in the results, as a strong genetic signal was found when performing classifications on this group. A combination of DNAm and SNPs were identified as the most informative at all time points, reflecting this homogeneity. The signal learnt during the cross-sectional experiments yielded impressive accuracies, F1 scores and improvements in accuracies with slightly higher metrics observed at years 1 and 2. This highlights that there is a robust signal contained in the integration of these modalities, which may be present even earlier in the disease course than what is identified here.

There is a strong preference in all models for including DNAm across all three experiment groups. DNAm was not included in the most predictive model in only one experiment for the idiopathic subgroup at year 0. Our findings show that the improvement in classification accuracy of DNAm is consistent across all time points. This prominence indicates that DNAm is predictive of PD at all time points of both subgroups. Considering the importance of DNAm in both genetic and idiopathic groups separately and combined suggests that there could be an overlapping signal contained in this modality. As DNAm is a measure of epigenetics, it suggests that there is common environmental or behavioural factors in both genetic and idiopathic groups which explains some aspect of their PD. Further research to identify the main drivers of variability in DNAm in the two subgroups separately and combined should be conducted to identify these factors.

Training a model that integrates SNPs and DNAm late in the disease course of individuals with a genetic predisposition for PD could form a viable early diagnostic tool. We obtain an average accuracy of 91% on a subset of participants in the PPMI dataset that have a genetic predisposition for PD and are present in either the SNPs or DNAm modality at each time point. Our results show that all models can accurately identify the HC class but, a model trained at year 3 is the best at distinguishing PD participants from PL at all time points. Training a model at year 3 is optimal, as the average disease state of a participant with PD will have progressed by this time. This makes it easier for the model to learn a threshold which can discriminate between PD and PL participants. This behaviour is evident from our model, as the accuracy achieved when predicting the PL class is robust when testing the model at earlier time points. Conversely, there is a deterioration in classification accuracy of the PD group when testing the model at earlier time points due to PD participants being misclassified as PL.

The most likely explanation of this phenomenon is that the effects of PD are not captured in all PD participants early in the disease course. As the SNPs dataset will differentiate perfectly between the HC class and the PD and PL genetic subgroups, this discriminatory effect is largely contained in the DNAm modality. DNAm is the process of binding methyl groups to sites in an individual’s DNA, resulting in alteration of expression [16]. It provides an epigenetic signature which can be inherited, associated with a disease and, depending on the site, reversed. Conditional to the DNA site affected, epigenetic modifications can occur slowly, meaning it can take a number of years for the effect of PD to be seen in a participant. In the PPMI study, DNAm was generated using whole-blood samples from participants. The advantage of using whole-blood samples is that they are minimally invasive and cost-effective. The disadvantage is that the biological signal may be quite weak for a neurological disorder in the blood due to the blood-brain barrier. This model also does not take into account individual participant trajectories. For example, two participants with PD may be recruited and diagnosed at the same time but can have different disease courses. This could further explain the decrease in accuracy at earlier time points of the PD class as some PD participants at these early time points may be at an earlier stage of the disease. Despite these limitations, we have shown excellent accuracy at all time points, making this a promising and viable approach to develop an early diagnostic tool for PD.

Diagnosing PD is a still an ongoing challenge of the disease, and being able to perform accurate early diagnosis would be a major step forward in the management of the disease. Diagnosis of PD in a clinical setting still involves the development of motor symptoms, by which time over 60% of dopamine neurons within specific regions of the basal ganglia may have been lost [17]. Pagan (2012) motivates that early detection can improve outcomes for PD patients by slowing disease progression and limiting its effect on their quality of life [17].

There are limitations to the model presented in this analysis. It is preferable that the sensitivity of the PD class rather than the specificity be accurate, as is the case here. If the sensitivity is high it means that the model is more likely to misdiagnose a PL participant as a PD which is preferable to misdiagnosing many PD participants. It cannot be determined how accurate this model is prior to a clinical PD diagnosis. This analysis is limited by the longitudinal time points of the PPMI dataset. Tracking the accuracy of this model for PL participants who go on to develop PD is a promising avenue of future research to further develop an early diagnostic tool. Further research also needs to be conducted in a dataset other than the PPMI dataset to measure the robustness of these findings. There is potential for survivor bias in the participants included in the longitudinal analysis. This analysis is limited by the use of a GCN. GCN is a transductive graph neural network algorithm, meaning all nodes have to be present during training and testing [9]). As a result, all participants are required to be present at each time point in order to be included in this longitudinal analysis. This leads to potential survivor bias, as all participants will have survived the disease until at least year 3 of this analysis. Future implementations should look towards inductive graph neural network algorithms which do not require all nodes to be present during training, thus allowing more participants to be included at each time point and eliminating survivor bias.

## V. Conclusion

This study highlights the importance of flexible integrative approaches to the analysis of PD. We have shown that there is a signal for PD present in genomic and proteomic data obtained from whole-blood samples. We have shown this both in a homogeneous group with a clear genetic driver for the disease and also in a more heterogeneous idiopathic group. We have achieved non-zero improvements in accuracy which are comparable to the MDS-UPDRS assessment baseline in the idiopathic group and significantly improved on this baseline in the genetic group. We have done so with models that do not account for the effects of medication or individual PD participant trajectory. We have identified DNAm as an informative omic measure in all individuals with PD and have proposed a model which could be used as an early diagnostic tool for individuals with a genetic predisposition for the disease. In summary, our research shows that an integrative network framework can be used to perform longitudinal stratification in PD.

## Supporting information

S1TablesFigures

## Data Availability

All data produced are available online at https://www.ppmi-info.org/access-data-specimens/download-data

## Author contributions statement

BR gathered all data, performed analysis, designed the study, conducted experiments and drafted the manuscript. TIS contributed to analysis, results and discussions. TIS and REM supervised the study, revised the manuscript and approved the final version of the manuscript.

## Acknowledgments

For the purpose of open access, the author has applied a creative commons’ attribution [CC BY] licence to any author accepted manuscript version arising.

Data used in the preparation of this article were obtained [on April, 5th 2022] from the Parkinson’s Progression Markers Initiative (PPMI) database (www.ppmi-info.org/access-dataspecimens/download-data), RRID:SCR 006431. For up-to-date information on the study, visit www.ppmi-info.org

PPMI – a public-private partnership – is funded by the Michael J. Fox Foundation for Parkinson’s Research and funding partners, including [list the full names of all the PPMI funding partners found on the PPMI Website].

## Code Availability

Code is available for download from : https://github.com/biomedicalinformaticsgroup/MOGDx-PPMI

## Supplementary Material

Supplementary figures and tables are available in S1FiguresTablesMOGDx.pdf

