## Supplementary material for "An Integrative Network Approach for Longitudinal Stratification in Parkinson’s Disease": S1TablesFigures

### Supplementary Figures

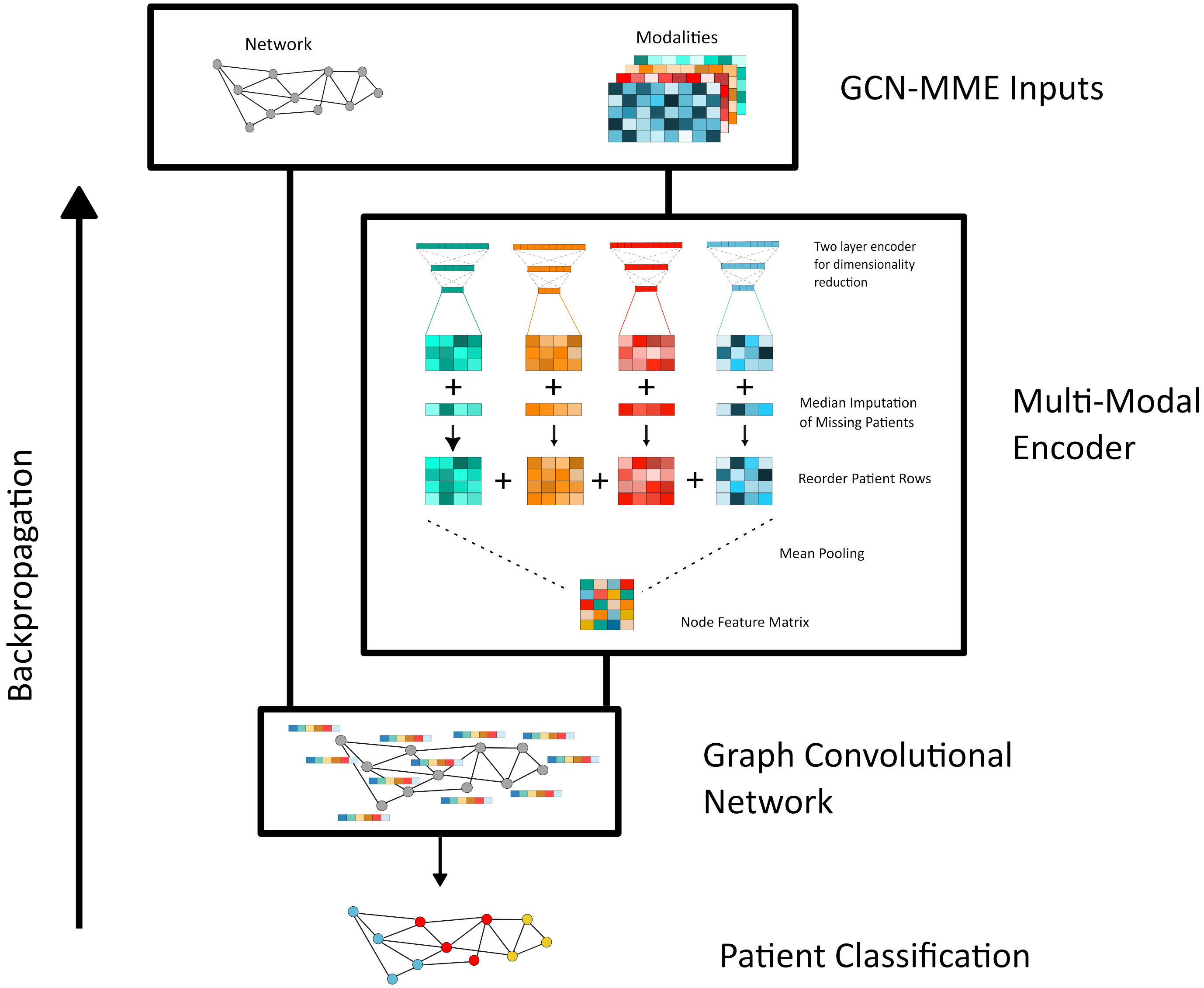


**Figure 1. Graph Convolutional Network - Multi-Modal Encoder Architecture (GCN-MME) |** The GCN-MME takes as input a fixed network and any number of modalities. The nodes in the network correspond to patients, and each patient is present in at least one modality. The modalities are encoded for dimensionality reduction using a two layer encoder. After the second layer, median imputation is performed to include patients missing from that modality but included in the network and at least one other modality. There is a shared latent embedding between the encoders, and the imputed second layers of each encoder are joined using mean pooling. This shared latent embedding forms the node features for the GCN. Patient classification is performed using the GCN with the loss back propagated through the entire GCN, thus, training each encoder in series with the GCN.

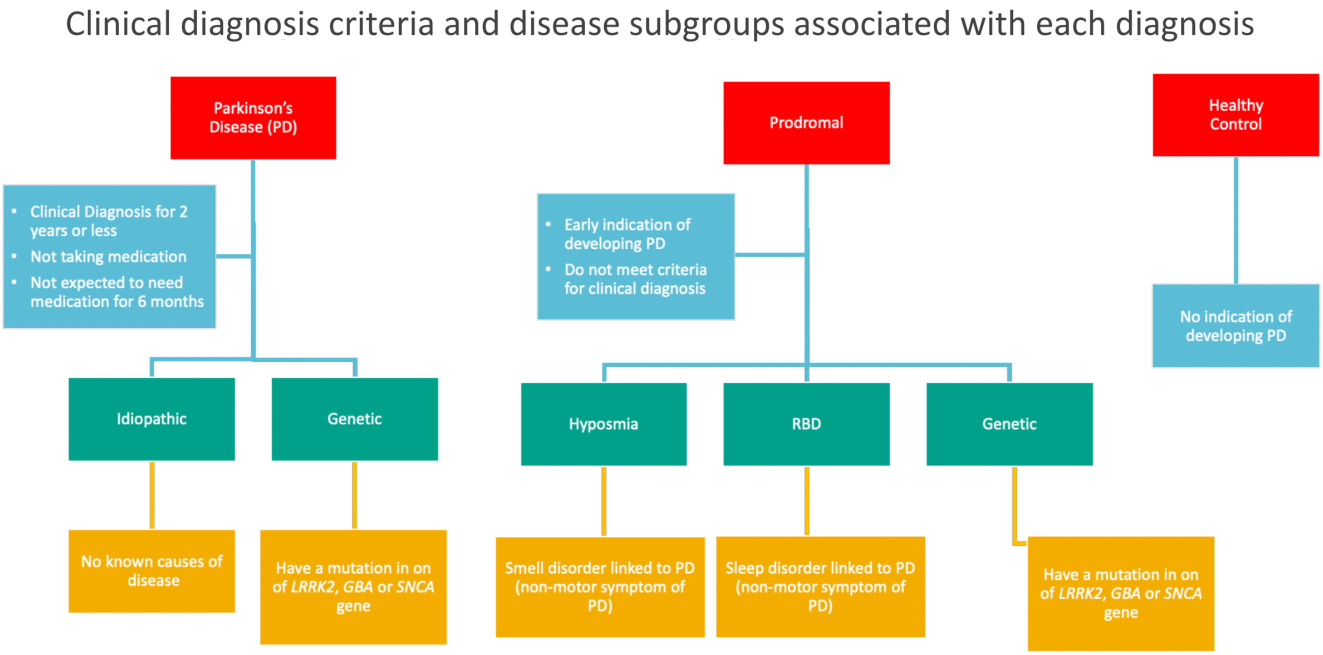


**Figure 2.** Breakdown of clinical diagnosis criteria and disease subtypes

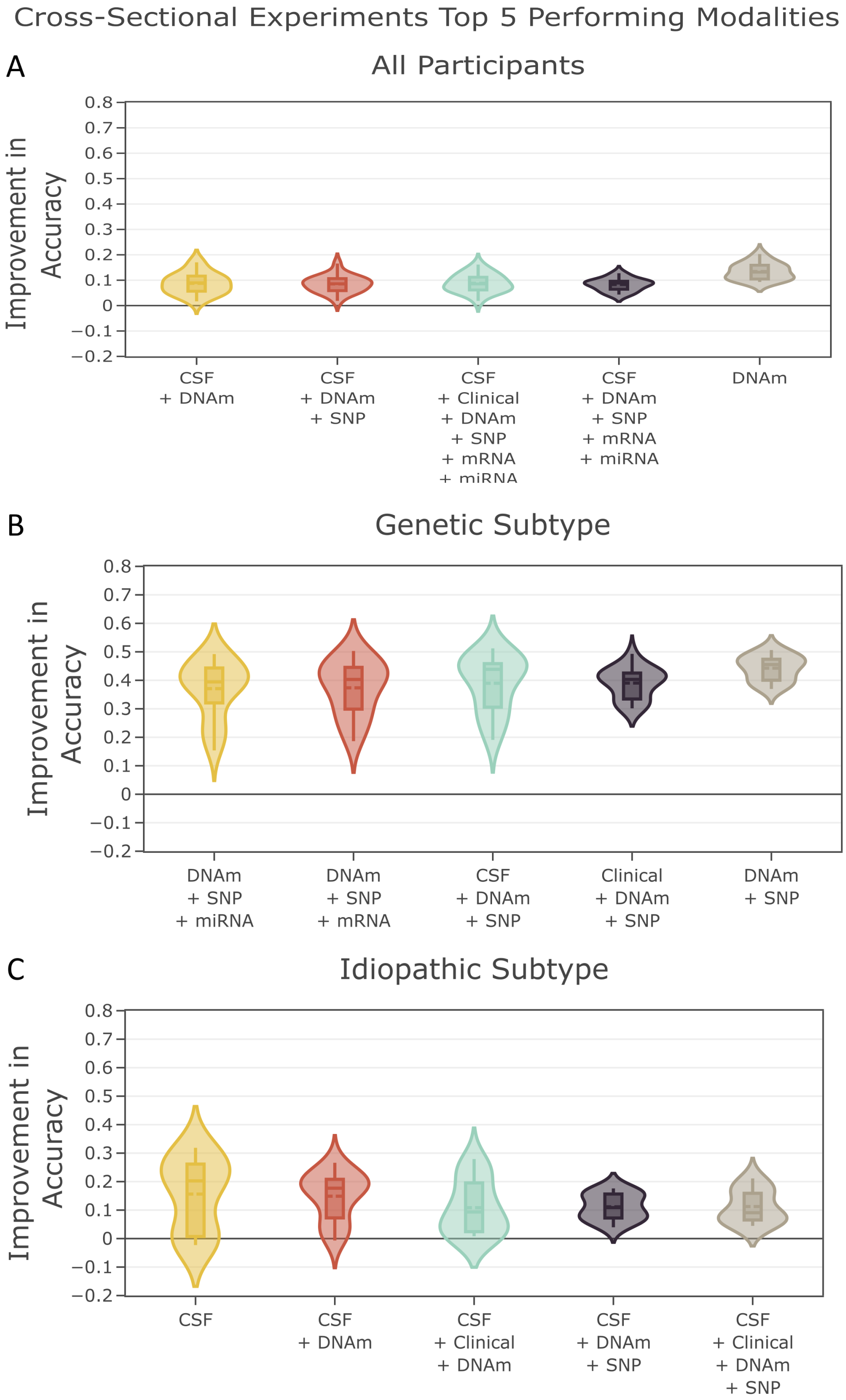


**Figure 3.** The top 5 modalities performing modalities in the cross-sectional experiments, averaged over time, and split by group

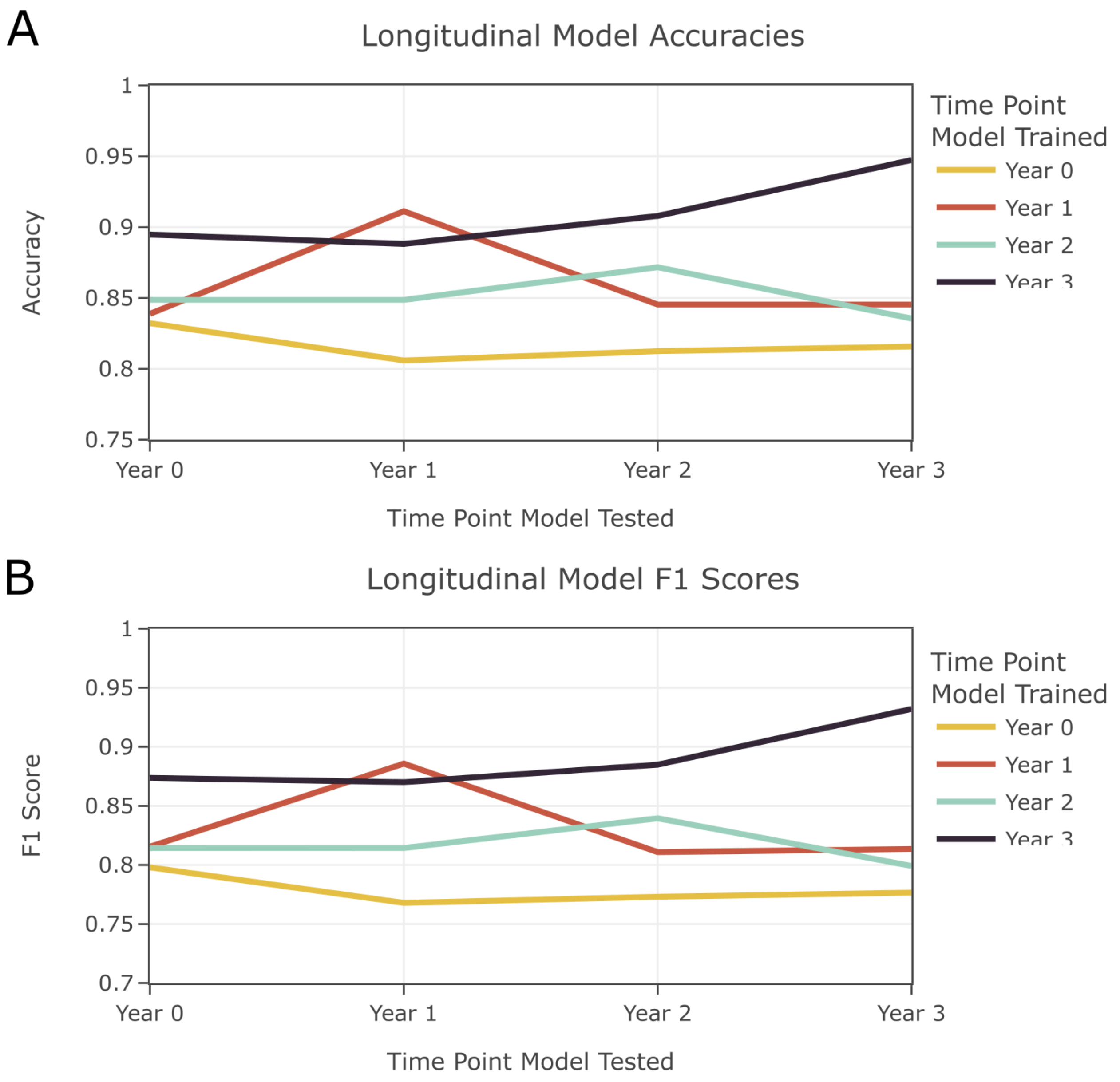


**Figure 4.** The performance metrics in the longitudinal experiment when models were trained at each time point and tested at all time points

### Supplementary Tables

**Table 1.** Breakdown of Modality Features in PPMI Dataset at Year 1

| | Raw Feature Count | Count After Processing | | | PSN Extracted Feature Count | | | Method of Extraction |
|---|---|---|---|---|---|---|---|---|
| | | All | Genetic | Idiopathic | All | Genetic | Idiopathic | |
| mRNA | 52338 | 26004 | 22158 | 34279 | 1143 | 1132 | 1143 | FDR <0.05 |
| miRNA | 40194 | 2730 | 2641 | 3094 | 329 | 396 | 406 | FDR <0.05 |
| DNAm | 805217 | 300k | 300k | 300k | 11 | 59 | 47 | $\lvert\omega\rvert > 0$ |
| SNP | 841 | 841 | 841 | 841 | 20 | 20 | 20 | None |
| Protein | 1472 | 1463 | 1463 | 1463 | 1463 | 1463 | 70 | $\lvert\omega\rvert > 0$ |
| Clinical | 6 | 6 | 6 | 6 | 6 | 6 | 6 | None |
| MDS-UPDRS | 88 | 64 | 64 | 64 | 32 | 39 | 14 | $\lvert\omega\rvert > 0$ |

**Table 2.** Breakdown of Modality Features in PPMI Dataset at Year 2

| | Raw Feature Count | Count After Processing | | | PSN Extracted Feature Count | | | Method of Extraction |
|---|---|---|---|---|---|---|---|---|
| | | All | Genetic | Idiopathic | All | Genetic | Idiopathic | |
| mRNA | 52338 | 27628 | 22736 | 37668 | 1101 | 1195 | 1146 | FDR <0.05 |
| miRNA | 40194 | 2987 | 3036 | 3190 | 307 | 513 | 434 | FDR <0.05 |
| DNAm | 805342 | 300k | 300k | 300k | 35 | 160 | 39 | $\lvert\omega\rvert > 0$ |
| SNP | 841 | 841 | 841 | 841 | 20 | 20 | 20 | None |
| Protein | 1472 | 1463 | 1463 | 1463 | 1463 | 1463 | 34 | $\lvert\omega\rvert > 0$ |
| Clinical | 6 | 6 | 6 | 6 | 6 | 6 | 6 | None |
| MDS-UPDRS | 88 | 64 | 64 | 64 | 22 | 2 | 25 | $\lvert\omega\rvert > 0$ |

**Table 3.** Breakdown of Modality Features in PPMI Dataset at Year 3

| | Raw Feature Count | Count After Processing | | | PSN Extracted Feature Count | | | Method of Extraction |
|---|---|---|---|---|---|---|---|---|
| | | All | Genetic | Idiopathic | All | Genetic | Idiopathic | |
| mRNA | 52338 | 39182 | 37866 | 41673 | 1150 | 889 | 1139 | FDR <0.05 |
| miRNA | 40194 | 3407 | 3175 | 3543 | 406 | 460 | 447 | FDR <0.05 |
| DNAm | 805106 | 300k | 300k | 300k | 300 | 20 | 77 | $\lvert\omega\rvert > 0$ |
| SNP | 841 | 841 | 841 | 841 | 20 | 20 | 20 | None |
| Protein | 1472 | 1463 | 1463 | 1463 | 1463 | 1463 | 13 | $\lvert\omega\rvert > 0$ |
| Clinical | 6 | 6 | 6 | 6 | 6 | 6 | 6 | None |
| MDS-UPDRS | 88 | 64 | 64 | 64 | 38 | 34 | 25 | $\lvert\omega\rvert > 0$ |

**Table 4.** Summary of PPMI Dataset by Sex, Visit and Subtype

| | Parkinson's Disease | | Prodromal | | | Healthy Control | Total |
|---|---|---|---|---|---|---|---|
| | Genetic | Sporadic | Genetic | RBD | Hyposmia | | |
| Participants | | | | | | | |
| Female | | | | | | | |
| <55 | 47 | 51 | 141 | 1 | 0 | 33 | 273 |
| 55-65 | 50 | 77 | 135 | 3 | 24 | 37 | 326 |
| >65 | 136 | 90 | 102 | 17 | 32 | 36 | 413 |
| Male | | | | | | | |
| <55 | 47 | 86 | 102 | 0 | 0 | 33 | 268 |
| 55-65 | 63 | 147 | 83 | 16 | 11 | 64 | 384 |
| >65 | 123 | 180 | 72 | 60 | 24 | 65 | 524 |
| Total | 466 | 631 | 635 | 97 | 91 | 268 | 2188 |
| Samples | | | | | | | |
| Female | | | | | | | |
| Year 0 | 233 | 218 | 378 | 21 | 56 | 105 | 1011 |
| Year 1 | 124 | 150 | 220 | 10 | 15 | 74 | 601 |
| Year 2 | 141 | 127 | 250 | 6 | 7 | 72 | 603 |
| Year 3 | 106 | 127 | 206 | 5 | 7 | 64 | 515 |
| Male | | | | | | | |
| Year 0 | 232 | 413 | 255 | 76 | 35 | 165 | 1176 |
| Year 1 | 120 | 298 | 161 | 40 | 20 | 129 | 768 |
| Year 2 | 141 | 238 | 173 | 30 | 15 | 108 | 705 |
| Year 3 | 92 | 233 | 142 | 28 | 12 | 102 | 609 |
| Total | 1189 | 1812 | 1785 | 216 | 167 | 819 | 5988 |

**Table 5.** Participant samples available per modality

| | | Parkinson's Disease | | Prodromal | | | Healthy Control |
|---|---|---|---|---|---|---|---|
| | | Idiopathic | Genetic | Genetic | RBD | Hyposmia | |
| mRNA | Year 0 | 364 | 327 | 417 | 27 | 20 | 190 |
| | Year 1 | 333 | 104 | 108 | 35 | 20 | 164 |
| | Year 2 | 318 | 116 | 84 | 36 | 21 | 157 |
| | Year 3 | 310 | 11 | 9 | 12 | 7 | 145 |
| miRNA | Year 0 | 370 | 305 | 357 | 35 | 23 | 179 |
| | Year 1 | 318 | 91 | 91 | 32 | 17 | 166 |
| | Year 2 | 327 | 99 | 70 | 34 | 15 | 156 |
| | Year 3 | 314 | 10 | 7 | 10 | 5 | 141 |
| DNAm | Year 0 | 223 | 88 | 131 | 29 | 21 | 190 |
| | Year 1 | 197 | 84 | 132 | 29 | 21 | 85 |
| | Year 2 | 204 | 82 | 127 | 28 | 19 | 82 |
| | Year 3 | 205 | 74 | 88 | 27 | 18 | 81 |
| SNP | Year 0 | 377 | 97 | 73 | 13 | 23 | 179 |
| | Year 1 | 331 | 71 | 60 | 12 | 21 | 173 |
| | Year 2 | 332 | 70 | 61 | 12 | 20 | 166 |
| | Year 3 | 330 | 64 | 56 | 11 | 17 | 158 |
| Protein | Year 0 | 399 | 206 | 351 | 0 | 0 | 182 |
| | Year 1 | 22 | 0 | 1 | 23 | 14 | 43 |
| | Year 2 | 43 | 0 | 1 | 15 | 5 | 72 |
| | Year 3 | 16 | 0 | 1 | 14 | 3 | 43 |
| Clinical | Year 0 | 223 | 88 | 131 | 29 | 21 | 86 |
| | Year 1 | 197 | 84 | 132 | 29 | 21 | 85 |
| | Year 2 | 204 | 82 | 127 | 28 | 19 | 82 |
| | Year 3 | 205 | 74 | 88 | 27 | 18 | 81 |
| MDS-UPDRS | Year 0 | 631 | 292 | 424 | 97 | 91 | 256 |
| | Year 1 | 452 | 243 | 380 | 50 | 35 | 202 |
| | Year 2 | 204 | 82 | 127 | 28 | 19 | 82 |
| | Year 3 | 360 | 183 | 314 | 33 | 19 | 165 |

**Table 6.** Subset of participants in longitudinal analysis

| | Genetic Predisposition | Control |
|---|---|---|
| Parkinson's Disease | 70 | 0 |
| Prodromal | 84 | 0 |
| Healthy Control | 0 | 150 |
| Total | 154 | 150 |

Participants in the genetic predisposition column have a mutation in one of their *LRRK2*, *SNCA* or *GBA* genes

**Table 7.** Longitudinal Experiments Performance Metrics

| | | Time point at which model was tested | | | | | | | |
|---|---|---|---|---|---|---|---|---|---|
| | | Year 0 | | Year 1 | | Year 2 | | Year 3 | |
| | | Accuracy | F1 | Accuracy | F1 | Accuracy | F1 | Accuracy | F1 |
| Time point at which model was trained | Year 0 | 0.832 | 0.798 | 0.806 | 0.768 | 0.813 | 0.773 | 0.816 | 0.777 |
| | Year 1 | 0.839 | 0.815 | 0.911 | 0.886 | 0.845 | 0.811 | 0.845 | 0.814 |
| | Year 2 | 0.849 | 0.814 | 0.849 | 0.814 | 0.872 | 0.840 | 0.836 | 0.799 |
| | Year 3 | 0.895 | 0.874 | 0.888 | 0.870 | 0.908 | 0.885 | 0.947 | 0.932 |
